# Evaluating Antibiotic Review Timing, Clinical Characteristics, and Stewardship Interventions to Confront Antimicrobial Resistance: A Retrospective Cohort Study of 640 Patients in Two UK Hospitals Before and During the COVID-19 Pandemic

**DOI:** 10.1101/2025.07.01.25330546

**Authors:** Rasha Abdelsalam Elshenawy, Nkiruka Umaru, Zoe Aslanpour

## Abstract

**Introduction:** Antimicrobial resistance (AMR) poses a critical global health threat, with projections of 10 million annual deaths by 2050 if left unaddressed. Antimicrobial stewardship (AMS) initiatives, such as the UKHSA *Start Smart – Then Focus* (SSTF) framework, are vital for optimising antibiotic use. However, the COVID-19 pandemic significantly disrupted AMS practices, leading to increased empirical prescribing and reduced opportunities for timely antibiotic review.

**Methods:** This retrospective cohort study analysed antibiotic review practices among 640 adult inpatients treated for respiratory tract infections (RTIs) at two secondary care hospitals within an English NHS Trust during 2019 (pre-pandemic) and 2020 (pandemic). Data included demographics, comorbidities, antibiotic classification (WHO AWaRe), review timing (Days 2–3, 4, 7), and AMS interventions aligned with SSTF-CARES outcomes. Statistical analysis was performed using SPSS v22.0.

**Results:** Patients were predominantly elderly (median age: 78–81), with high prevalence of comorbidities including hypertension (45%) and diabetes (20%). Overall mortality was 15%. Watch antibiotics were most frequently prescribed (46.3%–65.0%), especially during the pandemic, while Reserve antibiotic use remained appropriately low (<2%). Day 2–3 reviews accounted for 48.1%–54.4% of assessments, enabling timely AMS interventions such as de-escalation (up to 27.5%) and cessation (up to 35.6%). Hospital B consistently used more Watch antibiotics and performed more diagnostic imaging compared to Hospital A. Despite pandemic-related operational pressures, early reviews and stewardship activities were sustained.

**Conclusion:** Timely antibiotic review, particularly within 72 hours of initiation, supports effective stewardship and improves prescribing outcomes. The COVID-19 pandemic increased use of broad-spectrum agents but did not compromise early review practices. AMS resilience can be strengthened through standardised review protocols, institutional benchmarking, and integration of tools such as WHO’s AWaRe classification. Adaptive AMS strategies are essential for maintaining prescribing quality during public health emergencies.

**Trial Registration:** ISRCTN14825813

## Introduction

Antimicrobial resistance (AMR) is a growing global health emergency, predicted to cause up to 10 million deaths annually by 2050 if left unaddressed.^1^ In 2019, antibiotic-resistant infections directly caused 1.27 million deaths worldwide, while AMR was associated with 4.95 million deaths.^2–3^ At the forefront of efforts to mitigate AMR is antimicrobial stewardship (AMS)—an evidence-based, system-wide approach to promoting the judicious use of antimicrobials in order to optimise patient outcomes, reduce adverse effects, limit resistance, and preserve the efficacy of existing treatments.^3–7^

In the United Kingdom, where more than 90% of antibiotics are prescribed in primary and secondary care settings,^8^ the importance of AMS is underscored by national surveillance data. The 2024 English Surveillance Programme for Antimicrobial Utilisation and Resistance (ESPAUR) report documented a 3.5% increase in the AMR burden since 2019, largely driven by resistant Escherichia coli infections, and highlighted widening regional and ethnic disparities in prescribing.^9^

UK Health Security Agency’s (UKHSA) Start Smart – Then Focus (SSTF) is a national antimicrobial stewardship framework designed to support safe, timely, and appropriate antibiotic prescribing in inpatient settings.8 The framework operates on two key principles: “Start Smart” emphasises initiating the right antibiotic at the right dose for the right duration based on clinical assessment and local guidelines, while “Then Focus” involves systematic review and optimisation of antibiotic therapy based on clinical response and diagnostic results.^10^ A key element of this framework is the antibiotic review carried out 48–72 hours after initiating treatment. This allows clinicians to reassess the need for ongoing therapy using updated clinical and diagnostic information, in line with the “Then Focus” principle. Evidence shows that 20% of hospitalised patients experience antibiotic-related adverse events, underscoring the importance of timely reviews to minimise harm and optimise prescribing, as promoted by the UK’s Start Smart – Then Focus initiative.^11–13^

One critical AMS tool is antibiotic review, which audits prescribing practices to ensure appropriate antibiotic use, safety, and compliance with local guidelines.^14^ The timing of these reviews greatly influences their effectiveness, as different days focus on specific aspects: Day 1 reviews emphasise dose adjustments and IV-to-oral switches; Day 4 incorporates culture results to assess appropriateness; and Day 7 focuses on antibiotic therapy duration.^14–15^

The SSTF toolkit recommends applying the CARES outcomes—Cease, Amend, Refer, Extend, and Switch—to guide clinical decision-making.^16^ These structured antibiotic review outcomes support key stewardship interventions such as antibiotic stop, or discontinuing antibiotics when infection is unconfirmed, de-escalating to narrower spectrum agents, switching from intravenous to oral therapy, referring to specialist services when appropriate, or continuing antibiotic treatment with clearly defined review or stop dates. ^16–18^ This approach is central to optimising antibiotic use, improving patient outcomes, and reducing the development of antimicrobial resistance.^19^

The World Health Organisation’s AWaRe classification is a key tool in advancing AMS, helping to guide the appropriate use of antibiotics based on their risk of promoting resistance. It groups antibiotics into three categories: Access, Watch, and Reserve. Access to antibiotics is recommended for common infections due to their lower cost, narrower spectrum, and reduced potential to drive resistance. In contrast, Watch and Reserve antibiotics carry a higher risk and require careful monitoring.^20^ This system aligns with the UK’s Five-Year AMR Strategic Plan and supports coordinated international efforts to reduce antimicrobial resistance.^5^

The COVID-19 pandemic, however, exposed major weaknesses in AMS implementation.^21^ Despite WHO guidance discouraging antibiotic use without confirmed bacterial infection, antimicrobials were administered to approximately 70% of COVID-19 patients. This surge in inappropriate prescribing—particularly of Watch category antibiotics—risked accelerating resistance.^22–24^ Additional challenges, such as delays in microbiological testing, incomplete documentation, and limited multidisciplinary engagement, further disrupted timely antibiotic review and optimisation. These issues underline the urgent need to strengthen AMS systems, especially during public health emergencies.^25–27^

This retrospective cohort study aims to evaluate antibiotic review practices in two UK secondary care hospitals before and during the COVID-19 pandemic (2019–2020). It explores trends, clinical characteristics, and AMS interventions—particularly the adoption of CARES-aligned review decisions—to identify opportunities for strengthening stewardship resilience during health system disruptions.^5,28^

## Methods

### Study Design and Setting

This cross-sectional retrospective study was conducted at a single NHS Foundation Trust in England to evaluate antibiotic review practices, the timing of reviews (Days 2–3, 4, and 7), the appropriateness of antibiotic prescribing, and AMS interventions in adult patients with respiratory tract infections (RTIs) during 2019 (pre-pandemic) and 2020 (pandemic). The Trust includes two hospitals (A&B), serves a population of approximately 400,000 people, and has around 742 inpatient beds.

### Data Sources and Extraction

The primary author (RA) extracted data from the electronic medical records of patients within the Trust. Patient selection was based on electronic health record (EHR) entries identified by their respective International Classification of Diseases (ICD), ICD-10 codes for RTIs.^29^ This encompassed a range of conditions, including both specific and indeterminate diagnoses. Specific conditions included community-acquired pneumonia (CAP), chronic obstructive pulmonary disease (COPD), hospital-acquired pneumonia (HAP), and ventilator-associated pneumonia (VAP). Notably, in 2020, the selection also extended to cases of COVID-19 pneumonia. Alongside these, indeterminate diagnoses such as upper respiratory tract infections (URTIs), lower respiratory tract infections (LRTIs), and unspecified pneumonia were also considered. The primary diagnosis of RTIs in these records was pivotal in determining the initial or empirical antibiotic prescribed to the patients.

### Study Population and Eligibility

The study included adults aged 25 years and older, including pregnant and immunocompromised individuals, who were prescribed antibiotics for RTIs. Patients who spent less than 48–72 hours in the accident and emergency (A&E) department, were not prescribed antibiotics, or were under 25 years of age, were excluded. A stratified sampling approach ensured representation across demographic and clinical characteristics.

### Sample Size and Treated Population

A total of 640 patient records were reviewed and determined using Minitab software based on an estimated 20% inappropriate prescribing rate, which was calculated for a 10% margin of error and 95% confidence level. Records were evenly split between 2019 and 2020 (320 per year) and sampled across eight defined seasonal time points: March, June, September, and December of each year. Each time point included 80 patients, capturing both pre-pandemic and pandemic phases, including the first wave, national lockdown, second wave, and initial vaccination rollout.

### Clinical Characteristics and Variables

The data extraction captured demographics (age, gender), outcomes (discharge or death), allergy status, clinical indications, and comorbidities. Additional variables included timing of antibiotic review, prescribing decisions following review, chest X-ray findings, AWaRe classification (Access, Watch, Reserve), seasonal admission patterns, and AMS intervention types (e.g., change, continuation, de-escalation, escalation, IV-to-oral switch, cessation). Each record required approximately 45 minutes to review.

### Data Quality and Validation

The pilot test was essential to maintain the feasibility of the data extraction tool, ensure its validity and reliability, and support the integrity of the results and outcomes of the research project.^30^ In this research, a pilot study was conducted to provide an initial assessment of the dataset and to evaluate the feasibility of the data extraction tool in addressing the study’s research objectives. For validation, two independent authors extracted data from 1% of the sample (four patient records) separately, with a minimum agreement rate of 80% set as the benchmark for validity. To assess reliability, the same authors independently reviewed a second 1% sample. Inter-rater reliability was measured by calculating the percentage agreement between the independently extracted data. Any discrepancies were resolved through discussion to ensure consistency and accuracy.^31^

### Data Analysis

The study assessed antibiotic prescribing appropriateness according to local guidelines for both initial empirical selection (’Start Smart’) and subsequent decisions following clinical review (’Then Focus’). Data extraction captured demographics, outcomes, allergy status, clinical indications, and comorbidities. Antibiotic reviews were evaluated at specific intervals (Day 2-3, Day 4, and Day 7) to determine review timing and appropriateness of continued regimens, considering clinical response, microbiological results, and diagnostic findings like chest X-rays. Additional factors analysed included seasonal admission patterns, AWaRe classification (Access, Watch, Reserve), and types of antimicrobial stewardship interventions (change, continuation, de-escalation, escalation, IV-to-oral switch, cessation).

Descriptive statistics presented categorical variables as numbers and percentages, while non-normally distributed continuous variables were summarised using means and standard deviations. The prevalence of inappropriate prescribing was evaluated by comparing prescriptions to hospital antimicrobial guidelines during both pre-pandemic and pandemic periods. Implementation of antimicrobial stewardship was assessed using the UKHSA AMS Toolkit.^10^ Advanced statistical analyses were performed using IBM SPSS Statistics version 22.0.^32^

### Reporting and Ethical Considerations

The study followed the STROBE (Strengthening the Reporting of Observational Studies in Epidemiology) guidelines.^33^ The Health Research Authority (HRA) granted ethical approval for this study, with the Research Ethics Committee (REC) assigning reference number 22/EM/0161.^34^ In compliance with this approval, the study protocol underwent review and received approval from the University of Hertfordshire (UH) Ethics Committee under the reference LMS/PGR/NHS/02975.

### Patient and Public Involvement

The study protocol was submitted to the Citizens Senate, an organisation focused on patient care with a considerable representation of elderly individuals. They provided useful suggestions and comments.

### Study Registration

This study has been officially registered with the ISRCTN registry. The ISRCTN registry is a primary registry acknowledged by the WHO and the International Committee of Medical Journal Editors (ICMJE), accepting all clinical research studies.^35^ Additionally, it was registered in Octopus, the global primary research record.^36^

## Results

### Demographic and Patient Characteristics

The study included 640 participants (320 pre-pandemic, 320 during pandemic) with a median age of 78-81 years across all groups. The population was predominantly elderly, with 75% of patients aged ≥66 years. During the pandemic, there was a shift toward older patients, with those >85 years increasing from 24.4-29.3% in 2019 to 30.0-37.4% in 2020. Gender distribution remained balanced across all groups.

Overall mortality rates ranged from 13.8% to 16.2%, reflecting the high-risk nature of this population. Drug allergies were documented in 7.5-9.4% of patients across all groups. Community-acquired pneumonia was the most common indication for antibiotic therapy (33.1-45.0%), while COVID-19 emerged as a significant new indication in 2020 (10.6% Hospital A, 17.5% Hospital B). Hospital-acquired pneumonia showed variation between hospitals and time periods (11.9-23.1%).

For the comorbidities, hypertension was the most prevalent comorbidity (40.6-49.4%), followed by diabetes mellitus (15.6-22.5%) and kidney disease (13.8-23.8%). Notable pandemic-period changes included increased heart failure prevalence (particularly Hospital A: 11.3% to 21.3%), decreased kidney disease, and increased liver disease and depression rates. These findings reflect the complex, multimorbid population requiring antibiotic therapy in secondary care settings.

The combined data revealed 640 patients with a median age of 78 years (IQR 66-85), comprising 156 males (48.8%) and 164 females (51.2%). Overall mortality was 15.0% (48/320). Hypertension affected 143 patients (44.7%), diabetes mellitus 65 patients (20.3%), and kidney disease 75 patients (23.4%), confirming the substantial comorbidity burden.

**Table 1:** Demographic characteristics, clinical indications, comorbidities, and outcomes of patients with respiratory infections treated at two hospitals before (2019) and during (2020) the COVID-19 pandemic (Total Number is 640).

| Variable | Combined Data |  | Individual Hospital Data |  |  |  |
| --- | --- | --- | --- | --- | --- | --- |
|  | Prior to pandemic<br>(2019)<br>(n=320) | During pandemic<br>(2020)<br>(n=320) | Hospital A<br>2019<br>(n=160) | Hospital B<br>2019<br>(n=160) | Hospital A<br>2020<br>(n=160) | Hospital B<br>2020<br>(n=160) |
| **Age categories** |  |  |  |  |  |  |
| Age (Range), median (IQR) | 78 (66–85) | 79 (67–87) | 81 (73–88) | 80 (65–87) | — | — |
| • 25–45 years | 25 (7.8) | 19 (5.9) | 15 (9.4) | 10 (6.3) | 6 (3.8) | 13 (8.1) |
| • 46–65 years | 50 (15.6) | 48 (15.0) | 25 (15.6) | 25 (15.6) | 19 (11.9) | 29 (18.1) |
| • 66–85 years | 159 (49.7) | 145 (45.3) | 81 (50.6) | 78 (48.8) | 75 (46.9) | 70 (43.8) |
| • >85 years | 86 (26.9) | 108 (33.8) | 39 (24.4) | 47 (29.3) | 60 (37.4) | 48 (30.0) |
| **Gender** |  |  |  |  |  |  |
| • Male | 156 (48.8) | 165 (51.6) | 78 (48.8) | 78 (48.8) | 84 (52.5) | 81 (50.6) |
| • Female | 164 (51.2) | 155 (48.4) | 82 (51.2) | 82 (51.2) | 76 (47.5) | 79 (49.4) |
| **Patient outcome** |  |  |  |  |  |  |
| • Discharged | 272 (85.0) | 270 (84.4) | 138 (86.2) | 134 (83.8) | 134 (83.8) | 136 (85.0) |
| • Died | 48 (15.0) | 50 (15.6) | 22 (13.8) | 26 (16.2) | 26 (16.2) | 24 (15.0) |
| **Allergy** |  |  |  |  |  |  |
| • Allergy | 26 (8.1) | 27 (8.4) | 13 (8.1) | 13 (8.1) | 12 (7.5) | 15 (9.4) |
| • No allergy | 294 (91.9) | 293 (91.6) | 147 (91.9) | 147 (91.9) | 148 (92.5) | 145 (90.6) |
| **Clinical Indication** |  |  |  |  |  |  |
| • CAP | 126 (39.4) | 136 (42.5) | 53 (33.1) | 73 (45.6) | 69 (43.1) | 67 (41.9) |
| • COPD | 30 (9.4) | 14 (4.4) | 16 (10.0) | 14 (8.8) | 6 (3.8) | 8 (5.0) |
| • HAP | 67 (20.9) | 52 (16.2) | 40 (25.0) | 27 (16.9) | 33 (20.6) | 19 (11.9) |
| • VAP | 5 (1.6) | 1 (0.3) | 3 (1.9) | 2 (1.3) | 0 (0.0) | 1 (0.6) |
| • URTI | 6 (1.9) | 8 (2.5) | 6 (3.8) | 0 (0.0) | 0 (0.0) | 8 (5.0) |
| • LRTI | 30 (9.4) | 23 (7.2) | 12 (7.5) | 18 (11.3) | 9 (5.6) | 14 (8.8) |
| • Pneumonia | 56 (17.5) | 42 (13.1) | 30 (18.8) | 26 (16.3) | 24 (15.0) | 18 (11.3) |
| • COVID-19 | 0 (0.0) | 44 (13.8) | 0 (0.0) | 0 (0.0) | 19 (11.9) | 25 (15.6) |
| **Comorbidities** |  |  |  |  |  |  |
| • Hypertension (HPN) | 143 (44.7) | 148 (46.2) | 72 (45.0) | 71 (44.4) | 74 (46.3) | 74 (46.3) |
| • Hypotension | 13 (4.0) | 14 (4.4) | 7 (4.4) | 6 (3.8) | 7 (4.4) | 7 (4.4) |
| • Pulmonary embolism | — | — | 1 (0.6) | 2 (1.3) | 7 (4.4) | 7 (4.4) |
| • Atrial fibrillation | 61 (19.0) | 64 (20.0) | 31 (19.4) | 30 (18.8) | 32 (20.0) | 32 (20.0) |
|  | Prior to pandemic<br>(2019)<br>(n=320) | During pandemic<br>(2020)<br>(n=320) | Hospital A<br>2019<br>(n=160) | Hospital B<br>2019<br>(n=160) | Hospital A<br>2020<br>(n=160) | Hospital B<br>2020<br>(n=160) |
| • Heart failure | 32 (10.0) | 63 (19.6) | 16 (10.0) | 16 (10.0) | 32 (20.0) | 31 (19.4) |
| • Hypercholesteremia | 40 (12.5) | 58 (18.1) | 20 (12.5) | 20 (12.5) | 29 (18.1) | 29 (18.1) |
| • Diabetes mellitus | 65 (20.3) | 54 (16.9) | 33 (20.6) | 32 (20.0) | 27 (16.9) | 27 (16.9) |
| • Hypothyroidism | 24 (7.5) | 20 (6.2) | 12 (7.5) | 12 (7.5) | 10 (6.3) | 10 (6.3) |
| • Kidney disease | 75 (23.4) | 46 (14.4) | 38 (23.8) | 37 (23.1) | 23 (14.4) | 23 (14.4) |
| • Liver disease | 8 (2.5) | 19 (5.9) | 4 (2.5) | 4 (2.5) | 10 (6.3) | 9 (5.6) |
| • Malignancy | 50 (15.6) | 43 (13.4) | 25 (15.6) | 25 (15.6) | 22 (13.8) | 21 (13.1) |
| • Bronchiectasis | — | — | 3 (1.9) | 7 (4.4) | 20 (12.5) | 2 (1.3) |
| • Osteoarthritis | 31 (9.7) | 40 (12.5) | 16 (10.0) | 15 (9.4) | 20 (12.5) | 20 (12.5) |
| • Asthma | 35 (10.9) | 21 (6.5) | 18 (11.3) | 17 (10.6) | 11 (6.9) | 10 (6.3) |
| • COPD | 42 (13.1) | 40 (12.5) | 21 (13.1) | 21 (13.1) | 20 (12.5) | 20 (12.5) |
| • Dementia | 25 (7.8) | 23 (7.2) | 13 (8.1) | 12 (7.5) | 12 (7.5) | 11 (6.9) |
| • Epilepsy | 10 (3.1) | 13 (4.1) | 5 (3.1) | 5 (3.1) | 7 (4.4) | 6 (3.8) |
| • Depression | 12 (3.7) | 20 (6.2) | 6 (3.8) | 6 (3.8) | 10 (6.3) | 10 (6.3) |

**Note:** Data presented as n (%) unless otherwise specified. CAP = community-acquired pneumonia; COPD = chronic obstructive pulmonary disease; HAP = hospital-acquired pneumonia; VAP = ventilator-associated pneumonia; URTI = upper respiratory tract infection; LRTI = lower respiratory tract infection; HPN = hypertension. Age data is presented as the median (interquartile range). Comorbidities do not sum to 100% as patients may have multiple conditions. Missing values indicated by (—) where combined data not available or applicable.

### Antibiotic Classification and Diagnostic Imaging

With regards to the WHO AWaRe antibiotic classification system, Watch antibiotics constituted the predominant category of prescriptions across all study groups, accounting for 46.3% to 65.0% of total antibiotic use. Hospital B demonstrated consistently higher Watch antibiotic utilisation compared to Hospital A during both study periods (2019: 63.7% vs 46.3%; 2020: 65.0% vs 61.3%). A significant temporal shift occurred at Hospital A during the pandemic period, with prescribing patterns changing from predominantly Access antibiotics (51.9%) in 2019 to Watch antibiotics (61.3%) in 2020. Reserve antibiotics maintained appropriately low prevalence across all groups (0.0%-1.8%), indicating adherence to antimicrobial stewardship principles for last-resort agents.

For diagnostic imaging practices, chest radiography was not performed in the majority of patients across all study groups (52.5%-71.3%). Hospital A demonstrated consistently higher rates of non-imaging compared to Hospital B (70.0%-71.3% vs 52.5%-56.3% respectively). Among patients who underwent chest radiography, pneumonia was identified more frequently at Hospital B (18.1%-21.3%) compared to Hospital A (8.1%-10.6%). These observed patterns suggest institutional differences in clinical presentation characteristics, admission criteria, or established diagnostic protocols, with the pandemic period coinciding with increased utilisation of Watch category antibiotics across both hospitals.

**Table 2:** AWaRe antibiotic classification and diagnostic imaging results in patients treated at two hospitals before (2019) and during (2020) the COVID-19 pandemic (Total Number is 640).

| Variable | Hospital A 2019<br>(n=160) | Hospital B 2019<br>(n=160) | Hospital A 2020<br>(n=160) | Hospital B 2020<br>(n=160) |
| --- | --- | --- | --- | --- |
| **AWaRe Classification** |  |  |  |  |
| Access | 83 (51.8) | 57 (35.6) | 62 (38.8) | 54 (33.8) |
| Watch | 74 (46.3) | 102 (63.8) | 98 (61.2) | 104 (64.9) |
| Reserve | 3 (1.9) | 1 (0.6) | 0 (0.0) | 2 (1.3) |
| **Chest X-ray** |  |  |  |  |
| Pneumonia | 13 (8.1) | 29 (18.1) | 17 (10.6) | 34 (21.3) |
| No pneumonia | 35 (21.9) | 47 (29.4) | 29 (18.1) | 36 (22.5) |
| Not performed | 112 (70.0) | 84 (52.5) | 114 (71.3) | 90 (56.2) |

*Note: Data are presented as n (%). AWARe, Access, Watch, Reserve classification system for antibiotics as defined by the World Health Organisation.*

### Antibiotic Review Day Timing

Antibiotic review day timing demonstrated consistent patterns across both hospitals and time periods, with early reviews (Day 2-3) constituting the predominant practice. Day 2-3 reviews accounted for 48.1% to 54.4% of all antibiotic assessments, reflecting adherence to antimicrobial stewardship guidelines recommending early intervention. Hospital B showed higher rates of early review in 2019 compared to Hospital A (54.4% vs 48.8%), though this pattern reversed during the pandemic period (48.1% vs 53.8% respectively).

Day 4 reviews represented the second most common timing, ranging from 25.0% to 34.4% across groups. Hospital B demonstrated increased Day 4 review activity during the pandemic (34.4% vs 26.9% in 2019), while Hospital A showed a decrease (25.0% vs 31.3%). Day 7 reviews remained the least frequent category across all groups (17.5%-21.3%), with relatively stable patterns over time.

The predominance of early antibiotic reviews (Day 2-3 and Day 4 combined accounting for 73.1%-82.5% of all reviews) indicates effective implementation of antimicrobial stewardship protocols at both institutions. The temporal variations observed during the pandemic period may reflect adaptations to altered clinical workflows, increased clinical complexity, or modified review protocols in response to COVID-19 operational challenges.

### Antimicrobial Stewardship Interventions for Appropriate Prescribing

Among patients receiving appropriate antibiotic prescribing according to local antimicrobial guidelines, antimicrobial stewardship interventions demonstrated distinct patterns across hospitals and time periods (Figure 2). De-escalation to narrower-spectrum agents was the predominant intervention across all groups, accounting for the highest frequency of stewardship decisions. Hospital A showed consistent intervention activity with 68 total interventions in 2019 and 70 interventions in 2020, representing a slight increase during the pandemic period. Hospital B demonstrated lower overall intervention frequency, with 47 interventions in 2019 increasing to 61 interventions in 2020.

**Figure 1.**
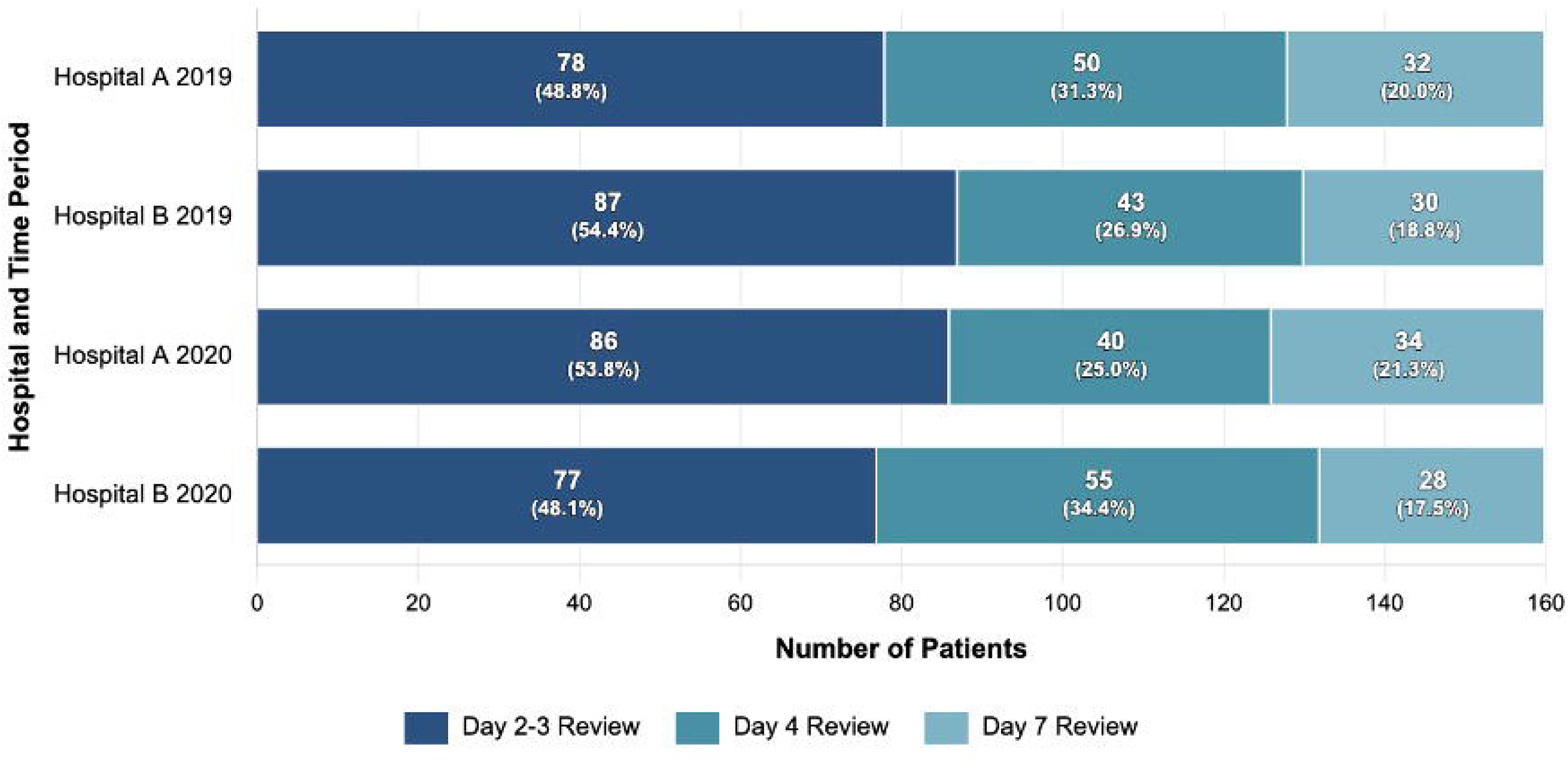
Antibiotic review day timing across study groups in two hospitals before (2019) and during (2020) the COVID-19 pandemic (Total Number is 640)

**Figure 2.**
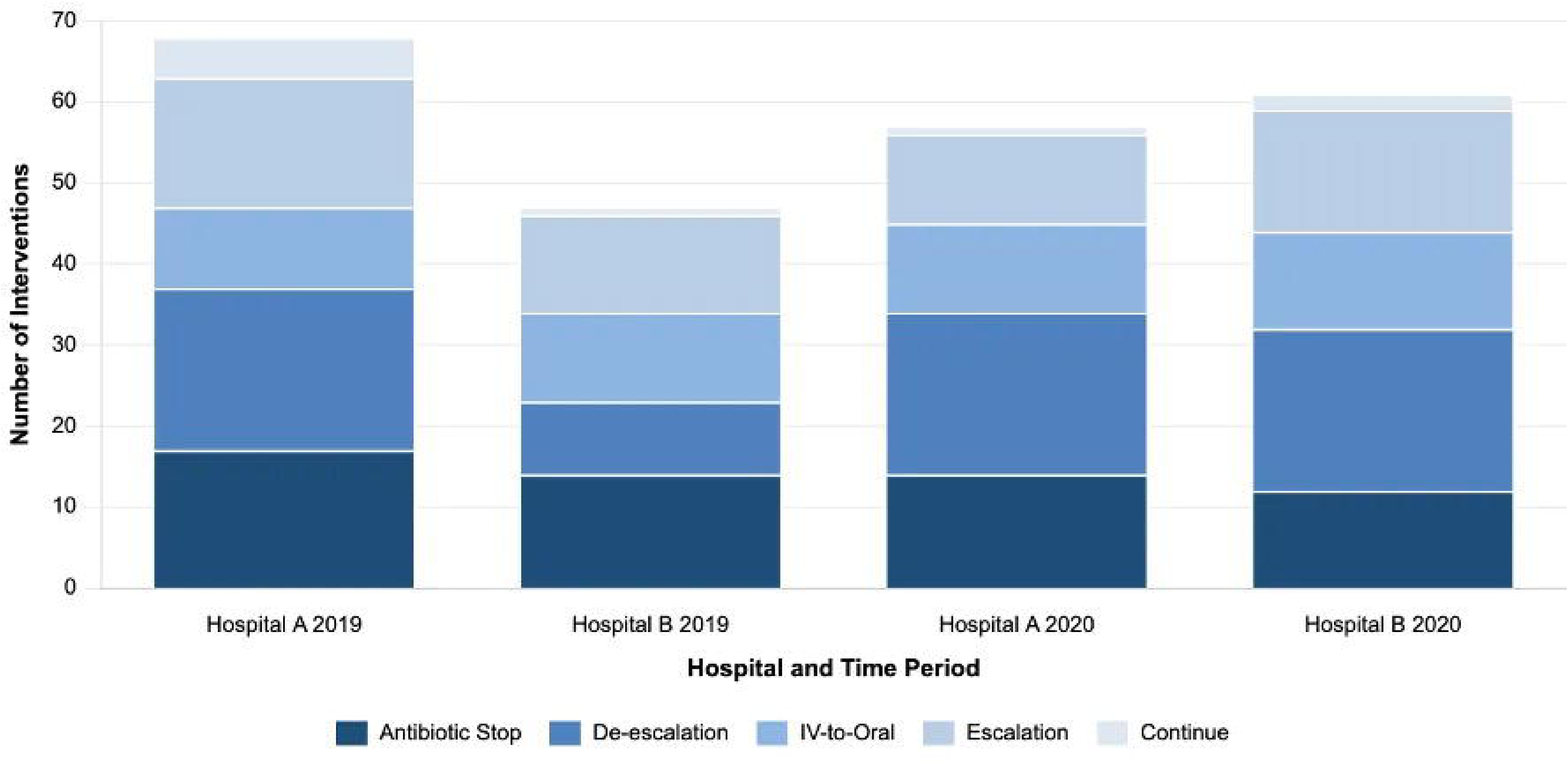
Antimicrobial Stewardship Interventions for Patients with Appropriate Antibiotic Prescribing Based on Local Guidelines in Two Hospital Settings Before (2019) and During (2020) the COVID-19 Pandemic.

Antibiotic discontinuation represented the second most common intervention, particularly at Hospital A during both time periods. Intravenous-to-oral conversion showed moderate activity across all groups, while escalation to broader-spectrum therapy occurred less frequently. Continuation of current antibiotic regimen was the least common intervention, suggesting active stewardship engagement rather than passive acceptance of existing prescriptions.

The increased intervention activity at both hospitals during the pandemic period may reflect heightened stewardship vigilance, altered patient acuity, or adaptive clinical practices in response to COVID-19. Despite initial appropriate antibiotic selection according to guidelines, substantial opportunities for further optimisation through stewardship interventions were identified, particularly de-escalation strategies that maintained therapeutic efficacy while reducing antimicrobial pressure.

## Discussion

This study examined antibiotic prescribing practices and antimicrobial stewardship interventions across two UK hospitals, comparing pre-pandemic (2019) and pandemic (2020) periods to evaluate antibiotic review timing,^14^ WHO AWaRe classification patterns,^20^ AMS Interventions,^10,27,28^ and patient characteristics influencing prescribing decisions. Our findings reveal critical insights into the evolving landscape of hospital antimicrobial use during a period of unprecedented healthcare challenges.

The demographic profile demonstrated a predominantly elderly population, with 75% of patients aged ≥66 years and median ages of 78-81 years across hospital groups, consistent with established literature showing higher antibiotic utilisation among older adults.^3,37^ During the pandemic, this ageing trend intensified, with patients >85 years increasing from 24.4- 29.3% to 30.0-37.4%, highlighting the urgent need for age-specific stewardship interventions addressing pharmacokinetic changes and increased adverse event susceptibility.^38^ Mortality rates of 13.8-16.2% aligned with NICE guidance for severe pneumonia outcomes.^39^, while community-acquired pneumonia’s predominance (33.1-45.0%) reflected its established role as the leading bacterial indication requiring hospitalisation.^40^

Regarding comorbidities, hypertension was most prevalent (44.7-46.3%), followed by diabetes mellitus (16.9-20.6%) and kidney disease (14.4-23.8%), patterns consistent with COVID-19 hospitalisation studies showing similar comorbidity distributions.^41,42^ Notably, the pandemic period revealed significant institutional variations, particularly Hospital A’s doubled heart failure rates (16.0% to 32.0%) and universal increases in depression (6.3-6.8% to 10.6-16.2%), reflecting the complex multimorbid burden requiring antibiotic therapy.

The combined data of 640 patients confirmed these trends, with consistent mortality (15.0%) and substantial comorbidity burden. These findings highlight the evolving complexity of patients requiring antimicrobial therapy and highlight the critical need for adaptive stewardship strategies that account for changing epidemiological patterns, demographic shifts, and the multifaceted clinical characteristics of modern hospital populations requiring antibiotic intervention.^43,46^

The AWaRe classification findings from this study reveal concerning patterns in antimicrobial prescribing practices that align with global trends but deviate from WHO targets. Our observation that Watch antibiotics predominated across all study groups (46.3%- 65.0%) contradicts the WHO’s 2023 target of achieving at least 60% Access antibiotic consumption globally. The WHO developed the AWaRe classification system as a tool to support antibiotic stewardship efforts at local, national and global levels, with antibiotics classified into Access, Watch and Reserve groups based on their impact on antimicrobial resistance.^20^ The marked institutional variation between hospitals, with Hospital B consistently demonstrating higher Watch antibiotic utilisation (63.7%-65.0% vs 46.3%- 61.3%), suggests significant differences in prescribing practices, local resistance patterns, or patient case-mix that warrant further investigation.

The pandemic-period shift at Hospital A from predominantly Access antibiotics (51.9%) to Watch antibiotics (61.3%) parallels findings from similar studies examining COVID-19’s impact on prescribing patterns. Research from an English NHS Foundation Trust examining 640 patients found that Watch category antibiotic use significantly increased during the COVID-19 pandemic in 2020, totalling 386 compared to 259 in the pre-pandemic year of 2019, with amoxicillin/clavulanic acid remaining the most frequently prescribed antibiotic. This convergence toward broader-spectrum agents may reflect clinician uncertainty regarding bacterial co-infection risks in COVID-19 patients, despite evidence suggesting low rates of true bacterial co-infection.^26^ The appropriately low Reserve antibiotic prevalence (0.0%- 1.8%) across all groups demonstrates adherence to antimicrobial stewardship principles for last-resort agents, indicating successful restriction policies for these critical antimicrobials.

The substantial variation in chest radiography practices between institutions (52.5%-71.3% non-imaging rates) raises important questions about diagnostic approaches and resource utilisation. Hospital A’s consistently higher non-imaging rates (70.0%-71.3%) compared to Hospital B (52.5%-56.3%) may reflect differences in clinical protocols, patient acuity, or institutional practices that could influence antibiotic prescribing decisions. These diagnostic imaging patterns, combined with the AWaRe prescribing data, suggest that institutional factors significantly influence antimicrobial stewardship outcomes and highlight the need for standardised approaches to optimise both diagnostic and therapeutic interventions.^47–49^

The predominance of early antibiotic reviews observed in our study aligns closely with established antimicrobial stewardship guidelines and contemporary literature. Evidence emphasises that diagnostic results typically become available 48-72 hours after initial presentation, providing the optimal window for antibiotic review and modification.^15^ This timing supports the UK’s “Start Smart, then Focus” strategy, which specifically promotes active review for all patients still on antibiotics 48 hours after admission, ^10,40^ corresponding to our findings where Day 2-3 reviews constituted 48.1-54.4% of all assessments. International guidance from the Centres for Disease Control and Prevention (CDC), the World Health Organisation (WHO), and other stewardship authorities consistently recommends antibiotic timeouts at 48-72 hours to reassess appropriateness when diagnostic information becomes available, supporting our observed patterns. ^17,14,28,45,50,51^

The British Society for Antimicrobial Chemotherapy (BSAC) *Practical Guide to Antimicrobial Stewardship* advocates for back-end stewardship strategies, which prioritise prospective audit and feedback at defined intervals.^14^ These approaches are generally more widely adopted and clinically accepted than restrictive front-end preauthorisation systems.^14^ The Singapore model exemplifies this framework, incorporating structured review points— such as Day 1 for IV-to-oral switch decisions, Day 4 for microbiology-guided therapy adjustments, and Day 7 for treatment duration reassessment.25 Our findings support this evidence-based model, demonstrating a pattern of early (Day 2–3) and intermediate (Day 4) antimicrobial reviews, consistent with the back-end strategy’s focus on post-prescription optimisation over initial prescribing restrictions.^28,52,53^

The effectiveness of structured antibiotic review timing is further validated by the Antibiotic Review Kit for Hospitals (ARK-Hospital) stepped-wedge cluster-randomised trial,^54^ which demonstrated that multifaceted interventions targeting antibiotic review decisions achieved sustained reductions in antibiotic use (−4.8% annually) without compromising patient safety.^54^ This aligns with our findings showing predominant early reviews (Day 2-3: 48.1-54.4%) followed by intermediate assessments (Day 4), supporting evidence-based review timing frameworks that optimise antibiotic stewardship through systematic post-prescription evaluation. Recent quality improvement studies have demonstrated significant challenges in achieving compliance with 48-hour review documentation, with baseline rates as low as 18% improving to 77% following targeted interventions, suggesting our hospitals performed relatively well with early review rates exceeding 48%.^55^ Prompt initiation of antibiotic therapy is vital for patient outcomes, but tailoring treatment based on microbiological data and clinical response is crucial. ^56^

Expert consensus recommends reviewing therapy within 24–72 hours to evaluate clinical progress and lab results, facilitating optimisation and potential de-escalation.^57^ This aligns with WHO AWaRe principles, prioritising Access antibiotics and limiting Watch and Reserve use to curb AMR, ensuring effective stewardship and improved patient safety.^26^ Current practice acknowledges that antibiotic durations are often arbitrary, with emerging approaches advocating for biomarker-guided therapy using inflammatory markers like C-reactive protein and procalcitonin to determine appropriate treatment endpoints rather than predetermined course lengths.^25^ The temporal variations observed during the pandemic period may reflect adaptations to altered clinical workflows, though our data suggests both institutions maintained adherence to evidence-based stewardship principles despite operational challenges.^58–59^

The distribution of antimicrobial stewardship interventions in our study reflects established patterns described in the literature and aligns with the UKHSA “Start Smart then Focus” framework.^10,27^ De-escalation emerged as the predominant intervention (18.1-27.5%), consistent with UKHSA guidance emphasising the review and revision of antimicrobials at 48-72 hours to switch from broad-spectrum to narrow-spectrum agents where appropriate. Antibiotic discontinuation represented the second most common intervention (20.6-35.6%), supporting the “cease” component of the CARES framework when no evidence of infection exists.^10,18,60^

The increased stewardship activity during the pandemic period, particularly de-escalation interventions, may reflect heightened antimicrobial awareness during COVID-19.^61^ This pattern aligns with recent literature demonstrating that structured stewardship programs can achieve significant intervention rates even among appropriately prescribed antibiotics according to local guidelines.^62–62^ The ARK-Hospital trial similarly found substantial opportunities for antibiotic optimisation through systematic review processes, with intervention rates comparable to our findings.^54^ The predominance of de-escalation and discontinuation interventions supports evidence that back-end stewardship strategies focusing on post-prescription review are more widely accepted and effective than restrictive front-end approaches, as recommended by current UKHSA guidance and international stewardship authorities.^14^ This study highlights the resilience of antimicrobial stewardship during the COVID-19 pandemic, with consistent early antibiotic reviews and active interventions across two hospitals. Our study highlights the need for adaptive antibiotic review timing to sustain robust AMS during healthcare crises. Early reviews (Day 2-3) enabled effective interventions like de-escalation and IV-to-oral switching, aligning with UKHSA’s framework. Despite increased Watch category prescribing during the pandemic, systematic reviews supported consistent stewardship. Future AMS programs must adopt innovative, flexible strategies to maintain effective antibiotic reviews, adapt to clinical demands, and ensure patient safety.

### Strengths and Limitations

This study offers several strengths. It analysed a large, representative sample of 640 patients from two UK secondary care hospitals, allowing comparisons across pre-pandemic and pandemic periods. Its design supports identification of temporal trends and pandemic-related disruptions to AMS. The study applied well-established national and global AMS frameworks, including the UKHSA Start Smart – Then Focus toolkit and WHO AWaRe classification, enabling benchmarking against best practice. A key strength is the detailed assessment of antibiotic review timing and stewardship interventions (e.g., de-escalation, cessation, IV-to-oral switch), providing practical insights for AMS implementation. Rigorous data quality measures, including piloting, inter-rater validation, and adherence to STROBE guidelines, further enhance credibility.

However, limitations include the retrospective design, which may introduce documentation bias and limit control of confounding variables. While conducted across two hospitals, both were within a single NHS Foundation Trust, which may limit broader generalisability.

Incomplete diagnostic imaging—particularly low chest radiograph use—may have influenced antibiotic decisions. The absence of microbiological culture data also limits assessment of appropriateness beyond guideline adherence. Finally, although seasonal sampling across four time points was applied, this approach may not fully reflect intra-annual variations in prescribing or stewardship activities. These factors should be considered when interpreting and applying the study’s findings.

## Conclusion

This study demonstrates the critical importance of adaptive antibiotic review timing in maintaining resilient antimicrobial stewardship during healthcare crises. Early reviews (Day 2-3) facilitated effective interventions, particularly de-escalation and IV-to-oral switching, supporting UKHSA’s “Start Smart-Then Focus” framework. Despite concerning increases in Watch category prescribing during the pandemic, systematic review processes enabled consistent stewardship interventions. Future AMS programmes must embrace innovative, flexible approaches to maintain robust antibiotic review practices while adapting to evolving clinical demands and preserving patient safety outcomes.

## Patient consent for publication

Not applicable.

## Ethics statements

This study received ethical approval from the Health Research Authority (HRA) with the Research Ethics Committee (REC) reference number 22/EM/0161. In addition, the study protocol was reviewed and approved by the University of Hertfordshire (UH) Ethics Committee under reference LMS/PGR/NHS/02975.

## Data Availability

All data produced in the present study are available upon reasonable request to the corresponding author, subject to approvals by the participating NHS Trust and relevant data governance policies.

## Acknowledgments

We thank the healthcare professionals and antimicrobial stewardship team at Bedfordshire Hospitals NHS Foundation Trust for their collaboration and support. We are particularly grateful to the pharmacy, microbiology, and Research & Development departments for their contributions to this study, and to the University of Hertfordshire, School of Life and Medical Sciences, for sponsoring the research.

## References

1 Price R. O’Neill report on antimicrobial resistance: funding for antimicrobial specialists should be improved. European Journal of Hospital Pharmacy 2016;23:245–7. 10.1136/ejhpharm-2016-001013

2 Antimicrobial Resistance Collaborators. Global Burden of Bacterial Antimicrobial Resistance in 2019: a Systematic Analysis. The Lancet 2022;399:629–55. 10.1016/S0140-6736(21)02724-0

3 Naghavi M, Vollset SE, Ikuta KS, et al. Global Burden of Bacterial Antimicrobial Resistance 1990–2021: a Systematic Analysis with Forecasts to 2050. The Lancet 2024;404:1199–226. 10.1016/s0140-6736(24)01867-1

4 National Institute for Health and Care Excellence (NICE). Antimicrobial stewardship: Systems and Processes for Effective Antimicrobial Medicine Use. Nice.org.uk. 2015.https://www.nice.org.uk/guidance/ng15

5 Department of Health and Social Care. Confronting antimicrobial resistance 2024 to 2029. GOV.UK. 2024.https://www.gov.uk/government/publications/uk-5-year-action-plan-for-antimicrobial-resistance-2024-to-2029/confronting-antimicrobial-resistance-2024-to-2029

6 Davey P, Marwick CA, Scott CL, et al. Interventions to improve antibiotic prescribing practices for hospital inpatients. Cochrane Database of Systematic Reviews 2017;2. 10.1002/14651858.cd003543.pub4

7 Agrawal S, Bapat A, Amos J, et al. Adopting prospective antimicrobial stewardship (AMS) practice in high-risk immunosuppressed groups: an urgent call to action in the era of antimicrobial resistance (AMR). JAC-Antimicrobial Resistance 2024;6. 10.1093/jacamr/dlae145

8 England N. NHS England» Digital vision for antimicrobial stewardship in England. England.nhs.uk. 2025.https://www.england.nhs.uk/long-read/digital-vision-for-antimicrobial-stewardship-in-england/

9 ESPAUR. ESPAUR report 2022 to 2023: lay summary. GOV.UK. 2024.https://www.gov.uk/government/publications/english-surveillance-programme-antimicrobial-utilisation-and-resistance-espaur-report/espaur-report-2022-to-2023-lay-summary

10 UKHSA. Start Smart Then Focus: antimicrobial stewardship toolkit for inpatient care settings. GOV.UK. 2024.https://www.gov.uk/government/publications/antimicrobial-stewardship-start-smart-then-focus/start-smart-then-focus-antimicrobial-stewardship-toolkit-for-inpatient-care-settings#section-2-sstf-principlesthen-focus

11 Tamma PD, Avdic E, Li DX, et al. Association of Adverse Events With Antibiotic Use in Hospitalized Patients. JAMA Internal Medicine 2017;177:1308. 10.1001/jamainternmed.2017.1938

12 Wojcik G, Ring N, Willis DS, et al. Improving Antibiotic Use in hospitals: Development of a Digital Antibiotic Review Tracking Toolkit (DARTT) Using the Behaviour Change Wheel. Psychology & Health 2023;1–21. 10.1080/08870446.2023.2182894

13 Selina Roy-Bentley, Neil Powell. Implementation of the Antimicrobial Review Kit (ARK) to optimise antimicrobial prescribing at the Royal Cornwall Hospital: a behavioural change odyssey. 2022.10.7861/clinmed.2021-0757

14 BSAC. Antimicrobial Stewardship - From Principles to Practice e-book. The British Society for Antimicrobial Chemotherapy. 2023.https://bsac.org.uk/antimicrobial-stewardship-from-principles-to-practice-e-book/

15 Cassim J, Essack SY, Chetty S. An audit of antibiotic prescriptions: an antimicrobial stewardship pre-implementation study at a tertiary care public hospital. JAC-Antimicrobial Resistance 2024;7. 10.1093/jacamr/dlae219

16 NHS England. NHS England» Antimicrobial resistance and antimicrobial stewardship pharmacy undergraduate competency framework. www.england.nhs.uk. 2024.https://www.england.nhs.uk/long-read/amr-and-ams-pharmacy-undergraduate-competency-framework/

17 Barlam TF, Cosgrove SE, Abbo LM, et al. Implementing an Antibiotic Stewardship Program: Guidelines by the Infectious Diseases Society of America and the Society for Healthcare Epidemiology of America. Clinical Infectious Diseases 2016;62:e51–77. 10.1093/cid/ciw118

18 De Waele JJ, Schouten J, Beovic B, et al. Antimicrobial de-escalation as part of antimicrobial stewardship in intensive care: no simple answers to simple questions—a viewpoint of experts. Intensive Care Medicine 2020;46:236–44. 10.1007/s00134-019-05871-z

19 Al-Omari A, Al Mutair A, Alhumaid S, et al. The impact of antimicrobial stewardship program implementation at four tertiary private hospitals: results of a five-years pre-post analysis. Antimicrobial Resistance & Infection Control 2020;9. 10.1186/s13756-020-00751-4

20 WHO. AWaRe classification of antibiotics for evaluation and monitoring of use, 2023. www.who.int. 2023.https://www.who.int/publications/i/item/WHO-MHP-HPS-EML-2023.04

21 World Health Organization. COVID-19 cases | WHO COVID-19 dashboard. WHO Data. 2025.https://data.who.int/dashboards/covid19/cases

22 Langford BJ, Soucy J-P .R., Leung V, et al. Antibiotic resistance associated with the COVID-19 pandemic: a systematic review and meta-analysis. Clinical Microbiology and Infection 2022;29. 10.1016/j.cmi.2022.12.006

23 WHO. Preventing the COVID-19 pandemic from causing an antibiotic resistance catastrophe. www.who.int. 2021.https://www.who.int/europe/news/item/18-11-2020-preventing-the-covid-19-pandemic-from-causing-an-antibiotic-resistance-catastrophe

24 Rasha Abdelsalam Elshenawy, Nkiruka Umaru, Amal Bandar Alharbi, et al. Antimicrobial stewardship implementation before and during the COVID-19 pandemic in the acute care settings: a systematic review. BMC Public Health 2023;23. 10.1186/s12889-023-15072-5

25 Ayodeji Matuluko, Valerie Ness, Jennifer Macdonald, Jacqueline Sneddon, Ronald Andrew Seaton, Kay Currie. The impact of the COVID-19 pandemic on the antimicrobial stewardship workforce in Scottish acute care hospitals—a qualitative study. 2024.10.1093/jacamr/dlae199

26 Abdelsalam Elshenawy R, Umaru N, Aslanpour Z. WHO AWaRe classification for antibiotic stewardship: tackling antimicrobial resistance - a descriptive study from an English NHS Foundation Trust prior to and during the COVID-19 pandemic. Frontiers in Microbiology 2023;14:1298858. 10.3389/fmicb.2023.1298858

27 Abdelsalam Elshenawy R, Umaru N, Aslanpour Z. Impact of COVID-19 on ‘Start Smart, Then Focus’ Antimicrobial Stewardship at One NHS Foundation Trust in England Prior to and during the Pandemic. COVID 2024;4:102–16. 10.3390/covid4010010

28 Chung GW, Wu JE, Yeo CL, et al. Antimicrobial stewardship. Virulence 2013;4:151–7. 10.4161/viru.21626

29 International Classification of Diseases (ICD). ICD-10 Version:2016. icd.who.int. 2020.https://icd.who.int/browse10/2016/en#/J00-J06

30 Rasha Abdelsalam Elshenawy, Nikkie Umaru, Aslanpour Z. Seasonal variations and the COVID-19 pandemic: impact on antimicrobial stewardship and antibiotic prescribing in a UK secondary care setting to combat antimicrobial resistance—a pilot study. Frontiers in Microbiology 2025;16:1–9. 10.3389/fmicb.2025.1530414

31 Piet Hanegraaf, Abrham Wondimu, Jacob Jan Mosselman, Rutger de Jong, Seye Abogunrin, Luisa Queiros, Marie Lane, Maarten J. Postma, Cornelis Boersma, Jurjen van der Schans. Inter-reviewer reliability of human literature reviewing and implications for the introduction of machine-assisted systematic reviews: a mixed-methods review. Bmj.com. 2023.doi: 10.1136/bmjopen-2023-076912

32 Advanced SPSS. Advanced Statistics - IBM SPSS Statistics. Ibm.com. 2024.https://www.ibm.com/products/spss-statistics/advanced-statistics

33 STROBE. STROBE – Strengthening the Reporting of Observational Studies in Epidemiology. STROBE. 2023.https://www.strobe-statement.org/

34 NHS Health Research Authority. NHS Health Research Authority. Health Research Authority. 2017.https://www.hra.nhs.uk/

35 ISRCTN. Antibiotic prescribing in an English secondary care setting before and during the COVID-19 pandemic. www.isrctn.com. 2022.https://www.isrctn.com/ISRCTN14825813

36 OCTOPUS. Evaluating Changes in Antibiotic Prescribing and AMS Practices at a UK NHS Trust: A Comparative Study of 2019 and the 2020 COVID-19 Period - Octopus | Built for Researchers. Octopusac Published Online First: 2019. 10.57874/pmsr-7s77

37 Luca Soraci, Cherubini A, Paoletti L, et al. Safety and Tolerability of Antimicrobial Agents in the Older Patient. Safety and Tolerability of Antimicrobial Agents in the Older Patient Published Online First: 28 March 2023. 10.1007/s40266-023-01019-3

38 Zhu Y, Liu Y, Jiang H. Geriatric Health Care During the COVID-19 Pandemic: Managing the Health Crisis. Clinical Interventions in Aging 2022;**Volume** 17:1365–78. 10.2147/cia.s376519

39 NICE. Summary of the evidence | Pneumonia (community-acquired): antimicrobial prescribing | Guidance | NICE. www.nice.org.uk. 2019.https://www.nice.org.uk/guidance/ng138/chapter/Summary-of-the-evidence

40 Ashiru-Oredope D, Sharland M, Charani E, et al. Improving the quality of antibiotic prescribing in the NHS by developing a new Antimicrobial Stewardship Programme: Start Smart--Then Focus. Journal of Antimicrobial Chemotherapy 2012;67:i51–63. 10.1093/jac/dks202

41 Bajgain KT, Badal S, Bajgain BB, et al. Prevalence of comorbidities among individuals with COVID-19: A rapid review of current literature. American Journal of Infection Control 2020;49. 10.1016/j.ajic.2020.06.213

42 Guan W, Liang W, Zhao Y, et al. Comorbidity and its impact on 1590 patients with Covid-19 in China: A Nationwide Analysis. European Respiratory Journal 2020;55:2000547. 10.1183/13993003.00547-2020

43 Tinker NJ, Foster RA, Webb BJ, et al. Interventions to optimize antimicrobial stewardship. Antimicrobial Stewardship & Healthcare Epidemiology 2021;1. 10.1017/ash.2021.210

44 Giamarellou H, Galani A, Karavasilis T, et al. Antimicrobial Stewardship in the Hospital Setting: A Narrative Review. Antibiotics 2023;12:1557–7. 10.3390/antibiotics12101557

45 CDC. Core Elements of Hospital Antibiotic Stewardship Programs. Antibiotic Prescribing and Use. 2024.https://www.cdc.gov/antibiotic-use/hcp/core-elements/hospital.html

46 Heil EL, Justo JA, Bork JT. Improving the Efficiency of Antimicrobial Stewardship Action in Acute Care Facilities. Open Forum Infectious Diseases 2023;10. 10.1093/ofid/ofad412

47 Khadse SN, Ugemuge S, Singh C. Impact of Antimicrobial Stewardship on Reducing Antimicrobial Resistance. Cureus 2023;15. 10.7759/cureus.49935

48 Zakhour J, Haddad SF, Kerbage A, et al. Diagnostic stewardship in infectious diseases: a continuum of antimicrobial stewardship in the fight against antimicrobial resistance. International Journal of Antimicrobial Agents 2023;62:106816. 10.1016/j.ijantimicag.2023.106816

49 Donà D, Barbieri E, Giulia Brigadoi, et al. State of the Art of Antimicrobial and Diagnostic Stewardship in Pediatric Setting. Antibiotics 2025;14:132–2. 10.3390/antibiotics14020132

50 WHO. Antimicrobial stewardship interventions: a practical guide. www.who.int. 2021.https://www.who.int/europe/publications/i/item/9789289056267

51 Ha DR, Haste NM, Gluckstein DP. The Role of Antibiotic Stewardship in Promoting Appropriate Antibiotic Use. American Journal of Lifestyle Medicine 2019;13:376–83. 10.1177/1559827617700824

52 Cheo Lian Yeo, Chan DW, Earnest A, et al. Prospective audit and feedback on antibiotic prescription in an adult hematology-oncology unit in Singapore. European Journal of Clinical Microbiology & Infectious Diseases 2012;31:583–90. 10.1007/s10096-011-1351-6

53 Elligsen M, Walker SAN, Pinto R, et al. Audit and Feedback to Reduce Broad-Spectrum Antibiotic Use among Intensive Care Unit Patients A Controlled Interrupted Time Series Analysis. Infection Control & Hospital Epidemiology 2012;33:354–61. 10.1086/664757

54 Llewelyn MJ, Budgell EP, Laskawiec-Szkonter M, et al. Antibiotic review kit for hospitals (ARK-Hospital): a stepped-wedge cluster-randomised controlled trial. The Lancet Infectious Diseases 2022;0. 10.1016/S1473-3099(22)00508-4

55 Endalamaw A, Khatri RB, Mengistu TS, et al. A scoping review of continuous quality improvement in healthcare system: Conceptualization, models and tools, barriers and facilitators, and impact. BMC health services research 2024;24:487. 10.1186/s12913-024-10828-0

56 Martínez ML, Plata-Menchaca EP, Ruiz-Rodríguez JC, et al. An approach to antibiotic treatment in patients with sepsis. Journal of Thoracic Disease 2020;12:1007–21. 10.21037/jtd.2020.01.47

57 King J, Chenoweth CE, England PC, et al. Early Recognition and Initial Management of Sepsis in Adult Patients. Ann Arbor (MI): Michigan Medicine University of Michigan 2023. https://www.ncbi.nlm.nih.gov/books/NBK598311/

58 Waldron C-A, Pallmann P, Schoenbuchner S, et al. Effectiveness of biomarker-guided duration of antibiotic treatment in children hospitalised with confirmed or suspected bacterial infection: the BATCH RCT. *Health technology assessment (Winchester*, England) 2025;29:1–125. 10.3310/MBVA3675

59 Nora D, Salluh J, Martin-Loeches I, et al. Biomarker-guided antibiotic therapy— strengths and limitations. Annals of Translational Medicine 2017;5:208–8. 10.21037/atm.2017.04.04

60 Gonzalez L, Cravoisy A, Barraud D, et al. Factors influencing the implementation of antibiotic de-escalation and impact of this strategy in critically ill patients. Critical Care 2013;17:R140. 10.1186/cc12819

61 Sullivan C, Fisher CR, Grabowsky L, et al. Combating Antimicrobial Resistance During the COVID-19 Pandemic: Perceived Risks and Protective Practices. Nih.gov. 2025.https://www.ncbi.nlm.nih.gov/books/NBK611796/

62 Yoon YK, Kwon KT, Jeong SJ, et al. Guidelines on Implementing Antimicrobial Stewardship Programs in Korea. Infection & Chemotherapy 2021;53:617–59. 10.3947/ic.2021.0098

63 Hulscher MEJL, Prins JM. Antibiotic stewardship: does it work in hospital practice? A review of the evidence base. Clinical Microbiology and Infection 2017;23:799–805. 10.1016/j.cmi.2017.07.017

